# Limited Cross-Variant Immunity after Infection with the SARS-CoV-2 Omicron Variant Without Vaccination

**DOI:** 10.1101/2022.01.13.22269243

**Authors:** Rahul K. Suryawanshi, Irene P Chen, Tongcui Ma, Abdullah M. Syed, Noah Brazer, Prachi Saldhi, Camille R Simoneau, Alison Ciling, Mir M. Khalid, Bharath Sreekumar, Pei-Yi Chen, G. Renuka Kumar, Mauricio Montano, Miguel A Garcia-Knight, Alicia Sotomayor-Gonzalez, Venice Servellita, Amelia Gliwa, Jenny Nguyen, Ines Silva, Bilal Milbes, Noah Kojima, Victoria Hess, Maria Shacreaw, Lauren Lopez, Matthew Brobeck, Fred Turner, Frank W Soveg, Ashley F. George, Xiaohui Fang, Mazharul Maishan, Michael Matthay, Warner C. Greene, Raul Andino, Lee Spraggon, Nadia R. Roan, Charles Y. Chiu, Jennifer Doudna, Melanie Ott

**Affiliations:** Gladstone Institutes, San Francisco, CA, USA; Biomedical Sciences Graduate Program, University of California San Francisco; San Francisco, CA, USA; Department of Medicine, University of California San Francisco; San Francisco, CA, USA; Quantitative Biosciences Institute COVID-19 Research Group, University of California San Francisco; San Francisco, CA, USA; Department of Microbiology and Immunology, University of California, San Francisco, San Francisco, CA, USA; Department of Laboratory Medicine, University of California, San Francisco, San Francisco, CA 94158, USA; Curative Inc., 430 S Cataract Ave San Dimas, CA, USA; Innovative Genomics Institute, University of California, Berkeley, Berkeley, CA, USA; Department of Molecular and Cell Biology, University of California, Berkeley, CA, USA; Molecular Biophysics and Integrated Bioimaging Division, Lawrence Berkeley National Laboratory, Berkeley, CA, USA; Howard Hughes Medical Institute, University of California, Berkeley, Berkeley, CA, USA; Department of Chemistry, University of California, Berkeley, Berkeley, CA, USA; California Institute for Quantitative Biosciences, University of California, Berkeley, Berkeley; Department of Urology, University of California, San Francisco, San Francisco, United States; Department of Medicine and Department of Anesthesia, Cardiovascular Research Institute, University of California San Francisco, San Francisco, CA, USA; UCSF-Abbott Viral Diagnostics and Discovery Center, San Francisco, CA, USA; Michael Hulton Center for HIV Cure Research at Gladstone; San Francisco, CA, USA

## Abstract

SARS-CoV-2 Delta and Omicron strains are the most globally relevant variants of concern (VOCs). While individuals infected with Delta are at risk to develop severe lung disease^1^, Omicron infection causes less severe disease, mostly upper respiratory symptoms^2,3^. The question arises whether rampant spread of Omicron could lead to mass immunization, accelerating the end of the pandemic. Here we show that infection with Delta, but not Omicron, induces broad immunity in mice. While sera from Omicron-infected mice only neutralize Omicron, sera from Delta-infected mice are broadly effective against Delta and other VOCs, including Omicron. This is not observed with the WA1 ancestral strain, although both WA1 and Delta elicited a highly pro-inflammatory cytokine response and replicated to similar titers in the respiratory tracts and lungs of infected mice as well as in human airway organoids. Pulmonary viral replication, pro-inflammatory cytokine expression, and overall disease progression are markedly reduced with Omicron infection. Analysis of human sera from Omicron and Delta breakthrough cases reveals effective cross-variant neutralization induced by both viruses in vaccinated individuals. Together, our results indicate that Omicron infection enhances preexisting immunity elicited by vaccines, but on its own may not induce broad, cross-neutralizing humoral immunity in unvaccinated individuals.

---

Since the beginning of the COVID-19 pandemic, multiple waves of infection have occurred from SARS-CoV-2 VOCs that continue to arise and out-compete preceding variants. VOCs with worldwide relevance are Delta (B.1.617.2) and most recently Omicron (B.1.1.529), while Alpha (B.1.1.7), Beta (B.1.351), and gamma (P.1) variants spread more locally. Delta and Omicron show an increasing number and complexity in spike mutations as well as mutations in other structural proteins, especially nucleocapsid, select nonstructural proteins, and accessory open reading frames. Omicron bears over 50 mutations across its genome, including some that overlap with previous variants but also novel ones, with 28 located within its spike glycoprotein.

The constellation of mutations in the Omicron spike protein has been associated with increased transmission^4^, decreased human angiotensin converting enzyme -2 (ACE2)-binding affinity^4^, decreased spike cleavage^5^, and decreased cell-to-cell fusion^5,6^. Importantly, Omicron spike mutations limit efficacies of neutralizing antibodies generated by previous infections, vaccines, and monoclonal antibody treatment^7^. Indeed, the risk of breakthrough infections and re-infections is increased with Omicron^8–10^. However, disease severity associated with Omicron is lower than Delta^10^, and prior infection or vaccination reduces the risk of hospitalization with Omicron^11,12^. Pressing questions are how effective Omicron-induced immunity is, whether it is cross-protective against other variants, and if prior infection with Delta confers protection against Omicron.

## Robust infection of mice and human airway cells by Delta and ancestral strains but not Omicron

To answer these questions, we studied WA1, Delta, and Omicron infections in mice. Because WA1 and Delta variants cannot infect regular laboratory mice^13^, we used transgenic mice overexpressing human ACE2 (K18-hACE2)^14^. We intranasally-infected these mice (10^4^ PFU) with the three viral isolates and subsequently monitored their body temperature and weight, which serves as indicators of disease progression over seven days (**Fig. 1a**). While Delta- and WA1-infected mice showed progressive hypothermia and severe weight loss during this time, Omicron-infected mice exhibited very mild symptoms with only a small increase in body temperature and no weight loss (**Fig. 1b,c**). Five days after infection, the WA1- and B.1.617.2-infected mice were hunched or lethargic, while the B.1.1.529-infected mice appeared completely normal (**Extended Data 1a**). All of the Omicron-infected mice survived the one-week experiment, while 100% of WA1- and 60% of the Delta-infected animals reached the humane end-point during this time (**Fig. 1e**). This replicates data previously obtained in infected individuals, mice, and hamsters that show mild disease with Omicron, but not with Delta and WA1 infections^2,15^.

**Fig. 1.**
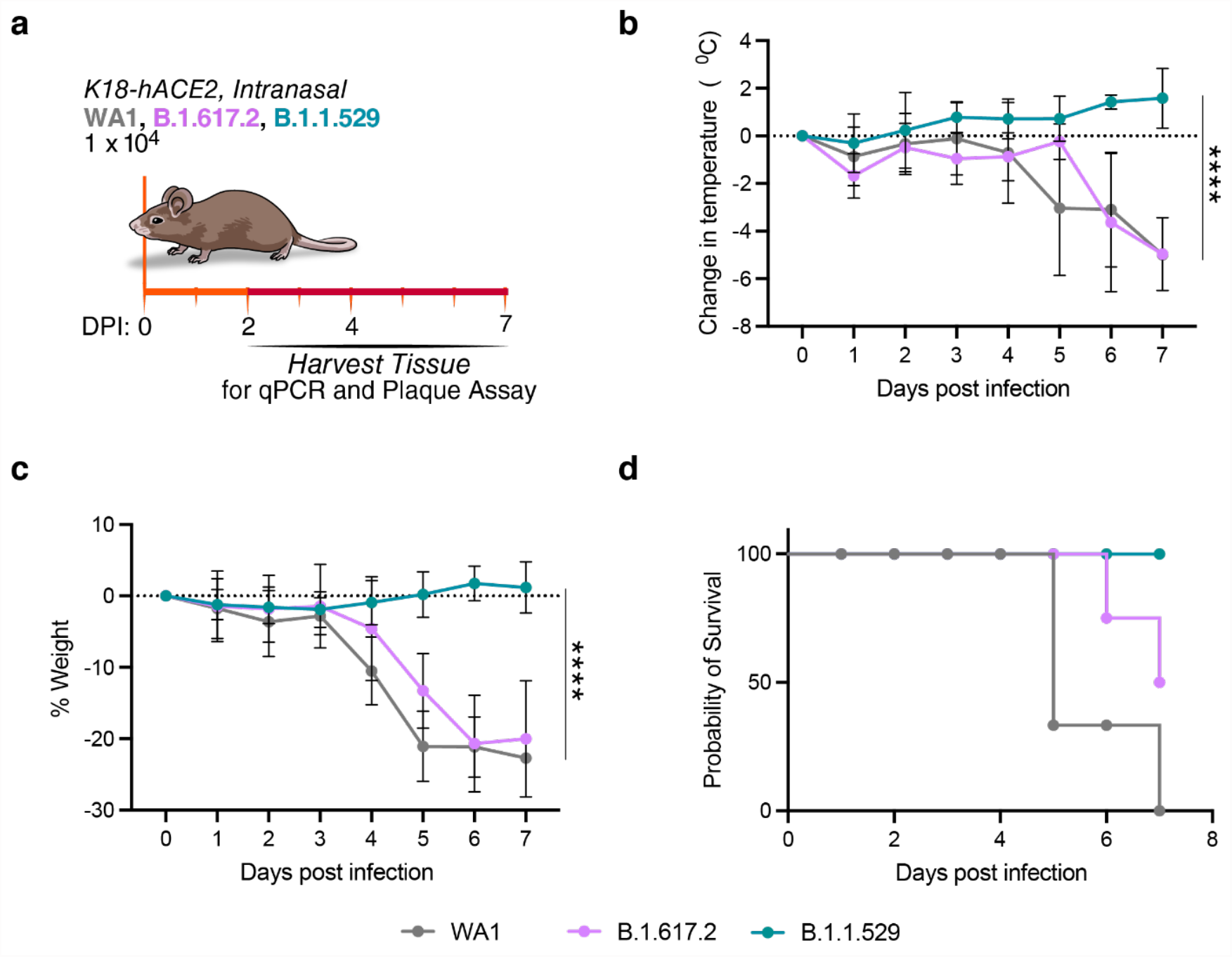
Robust infection of K18-hACE2 mice with B.1.617.2 and ancestral strain, but not B.1.1.529. **a**, Schematic of the experiment. 15 mice per group were intranasally infected with 10^4^ PFU of the indicated variant. Body temperature and weight were monitored daily. At the 2, 4, and 7 days post infection (dpi), the upper respiratory tract and lungs were harvested and processed for downstream analysis. **b**,Changes in body temperature of WA1 (grey), B.1.617.2 (purple), and B.1.1.529 (teal) infected mice. Data is shown as the average ± SD and analyzed by 2way ANOVA. ****p<0.0001. **c**, Severe weight loss of WA1- and B.1.617.2-infected mice. Data is shown as the average ± SD and analyzed by 2way ANOVA. ****p<0.0001. **d**, Probability of survival of variant infected mice.

To assess viral replication dynamics, we quantified infectious particle production (**Fig. 2a,b**) and viral RNA expression (**Extended Data 2A, B**) in the respiratory tracts and lungs of infected mice over time. Across all time points, high viral titers were present in airways and lungs of WA1-and Delta-infected mice, whereas Omicron replication was significantly lower (up to 3 logs) in these organs. In addition, brain tissue, which is a target for viral replication in K18-hACE2 mice^16^, showed lower Omicron replication at four and seven days after infection. Omicron also produced less infectious particles in human airway organoids and in the human alveolar A549 epithelial cell line overexpressing ACE2 relative to WA1 and Delta infections, consistent with the model that Omicron does not readily infect lower airway epithelial cells (**Fig. 2c,d and Extended Data 2e**). Notably we did not observe high Omicron titers in our respiratory tract samples (nasal turbinates and bronchi) in contrast to what others have recently observed in mouse nasal washes and human primary nasal and bronchial epithelial cells^2,17,18^. However, Omicron infectious titers decreased less over time in our respiratory tract samples as compared to the lungs, suggesting a more sustained replication of Omicron in this compartment (**Fig. 2a,b**).

**Fig. 2.**
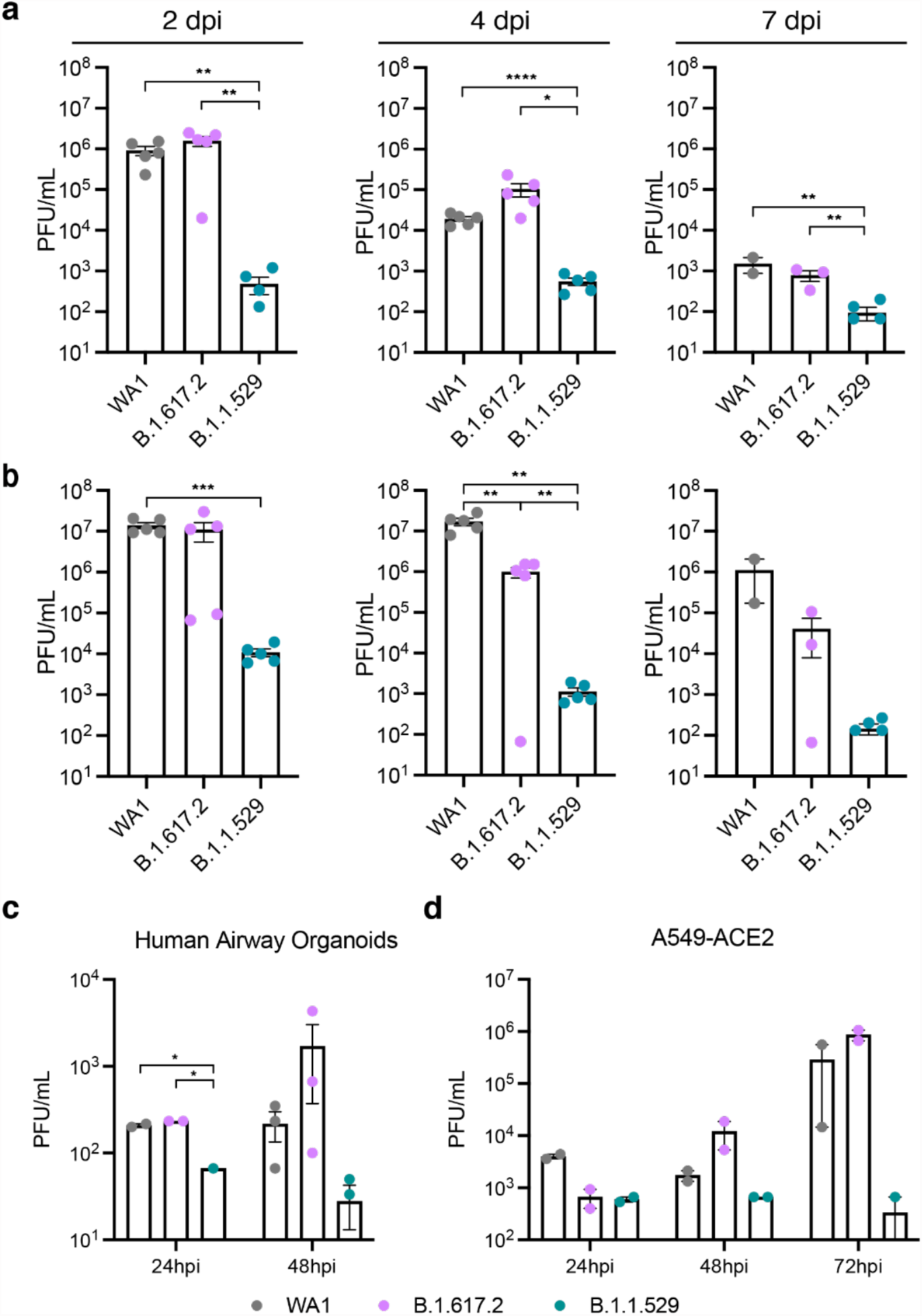
Robust viral replication of WA1 and B.1.617.2, but not B.1.1.529, in mice and human airway cells. **a**, Plaque assay titers from the upper airway (nasal turbinates and bronchus) of WA1 (grey), B.1.617.2 (purple), and B.1.1.529 (teal) infected mice at the indicated time point. Data is shown as the average ± SEM at 2, 4, and 7 dpi analyzed by the student’s t-test. *p<0.05, **p< 0.01, ****p<0.0001. **b**, Plaque assay titers from the lungs of infected mice at the indicated time point. Data is shown as the average ± SEM at each time point and analyzed by student’s t-test. **p< 0.01, ***p=0.0005. **c**, Plaque assay titers from supernatants of infected human airway organoids (MOI of 1). Data is shown as the average ± SEM and analyzed by 2wayANOVA. *p<0.05. **d**, Plaque assay titers from supernatants of infected A549-ACE2 cells (Multiplicity of infection [MOI] of 0.1). Data is shown as the average ± SEM.

## Inflammatory and immune markers differ between variants

Next, we measured mRNA expression of inflammatory and innate immune parameters in infected mouse lungs. While infection with WA1 and Delta readily induced proinflammatory markers of severe COVID, such as CXCL10 and CCL2^19^, induction by Omicron was significantly reduced early after infection (**Fig. 3a**). Although induction of interleukin 1α (IL1α) was not significantly different between the three viral strains, it trended towards lower expression in Omicron-infected animals two days post-infection (**Fig. 3a**). No significant differences were observed in the induction of interferon-α (IFNα) or relevant downstream induced genes such as interferon-stimulated gene 15 (ISG15) and 2’-5’-oligoadenylate synthetase 1 (OAS1), which was surprising given the markedly lower viral replication of the Omicron strain (**Fig. 3b**). These results show lower expression of some proinflammatory genes but induction of innate antiviral immunity in the lungs of Omicron-infected mice, which could explain the mild disease phenotype observed in these mice.

**Fig. 3.**
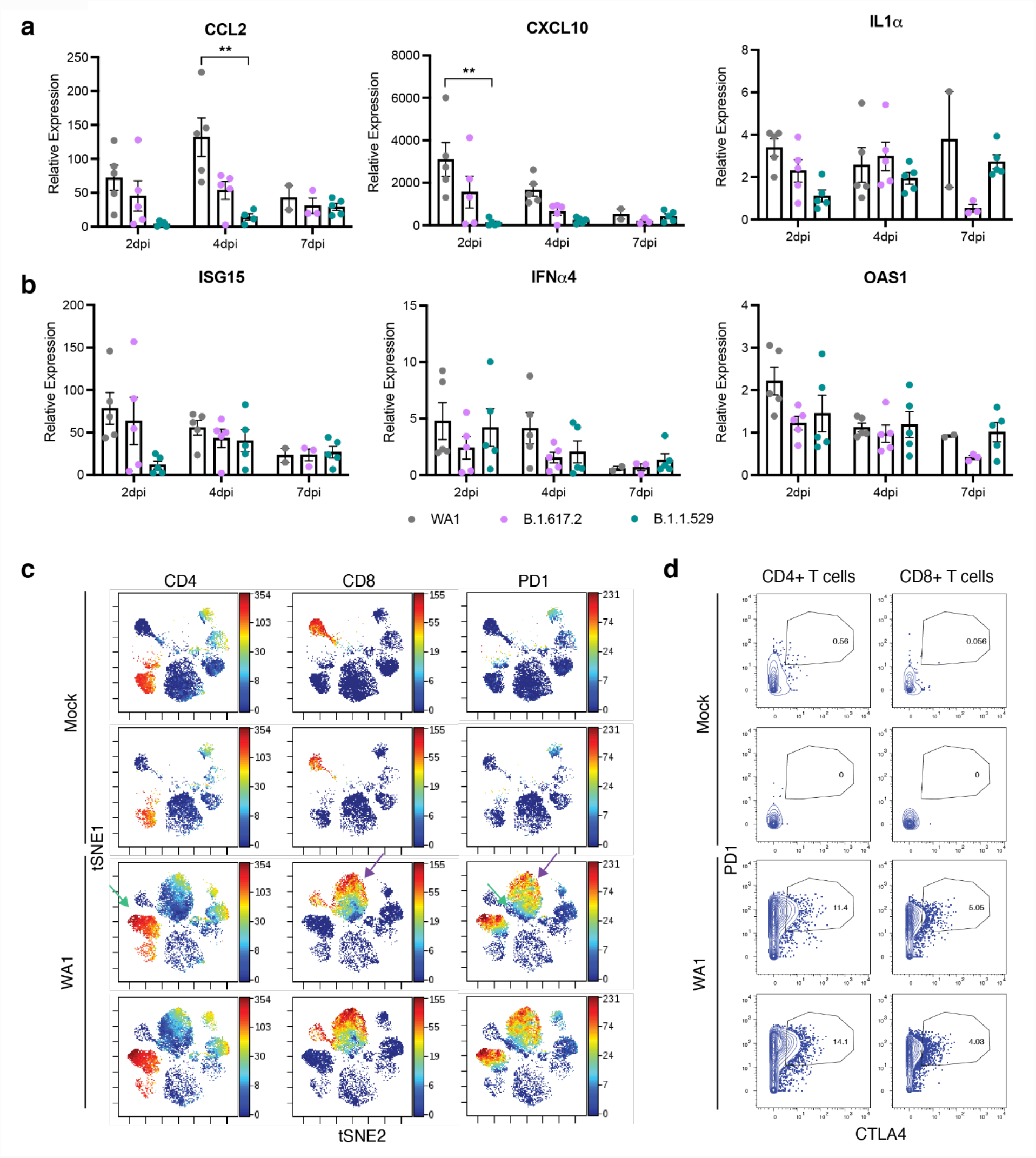
Differential expression of proinflammatory markers in lungs of infected mice and account of SARS-CoV-2 specific T-cell response. **a**, RT-qPCR of proinflammatory cytokines and chemokines from RNA isolated from lungs of infected mice at the indicated time points. Data are expressed relative to mock infected mice. Data is shown as the average ± SEM and analyzed by student’s t-test. **p<0.001. **b**, RT-qPCR of interferon stimulated genes from RNA isolated from lungs of infected mice at the indicated time points. Data are expressed relative to mock infected mice. Data is shown as the average ± SEM and analyzed by student’s t-test. **c**, T cells from lungs of infected mice are phenotypically distinct and express PD1. Single-cell suspensions of lungs from two mock infected (top two rows) and two WA1-infected (bottom two rows) K18-hACE2 mice were harvested 9 dpi and then analyzed by CyTOF. Shown are tSNE plots gated on total immune cells (CD45+) from the lungs of the mice, colored by expression levels of the antigen listed at the top (red = highest expression, blue = lowest expression). “Islands” of CD4+ and CD8+ T cells unique to the infected mice (identified by the green and purple arrows, respectively, in the third row) express especially high levels of the activation/exhaustion marker PD1, as demonstrated in the right-hand column. **d**, T cells from lungs of infected mice preferentially co-express the exhaustion markers PD1 and CTLA4. The T cells depicted in panel B were gated for PD1 and CTLA4 expression levels and depicted as 2D plots.

As severe COVID-19 is associated with cytokine storms in conjunction with exhaustion of T cells^20^, we next assessed whether the highly pro-inflammatory response we observed in WA1-infected mice is also associated with T cell exhaustion in late infection. We generated single-cell suspensions from the lungs of mock- and WA1-infected mice (9 dpi), and performed Cytometry by Time of Flight (CyTOF) mass spectrometry before and after stimulation with overlapping 15-mer peptides spanning the entire spike protein. tSNE visualization of the CyTOF data corresponding to total immune (CD45+) cells from the unstimulated specimens revealed that both CD4+ and CD8+ T cells of infected mice segregate distinctly from their respective counterparts in the mock-infected mice, indicating profound phenotypic changes in pulmonary T cells upon WA1 infection (**Fig. 3c**). One notable feature of the T cells from the infected mice was the high expression of the exhaustion markers programmed cell death 1 (PD1) and cytotoxic T-lymphocyte-associated protein 4 (CTLA4), which by contrast were barely expressed on T cells from mock-infected mice (**Fig. 3c, Extended Data 3a**). Despite this severe state of exhaustion, however, functional SARS-CoV-2-specific T cells were still primed, as demonstrated by our identification of IFNγ- and TNFα-producing cells specifically in the peptide-stimulated specimens (**Extended Data 3b**). These results suggest that WA1 infection elicits a highly pro-inflammatory cytokine response and exhausted pulmonary T cells in K18-hACE2 mice.

## Infection with Delta, but not Omicron, induces cross-variant neutralization

To determine humoral immune responses induced by infection with the three different strains, we collected sera from infected mice and tested their neutralization efficiency against SARS-CoV-2 strains: WA1, Alpha, Beta, Delta, and Omicron. Efficient virus neutralization was defined by more than 50% reduction in plaque forming units at the lowest serum dilutions (1:5 and 1:15)^21,22^. We also calculated the 50% neutralization titers (NT50) of the individual sera against the different viral strains (**Table 1**). Sera from Delta-infected mice showed the broadest cross-variant neutralization, effectively neutralizing all the strains except Beta (**Fig. 4b,d, Table 1**). By contrast, while Omicron infection effectively neutralized Omicron itself, it exhibited limited (<50%) cross-neutralization of other strains (**Fig. 4c,d, Table 1**). Sera from WA1-infected mice conferred effective protection against WA1, Alpha, and Delta, but not against Beta and Omicron (**Fig. 4a,d, Table 1**). These results indicate limited immunity induced by Omicron relative to other strains, which may be due to its highly mutated spike protein or its lower replicative capacity.

**Table 1.**
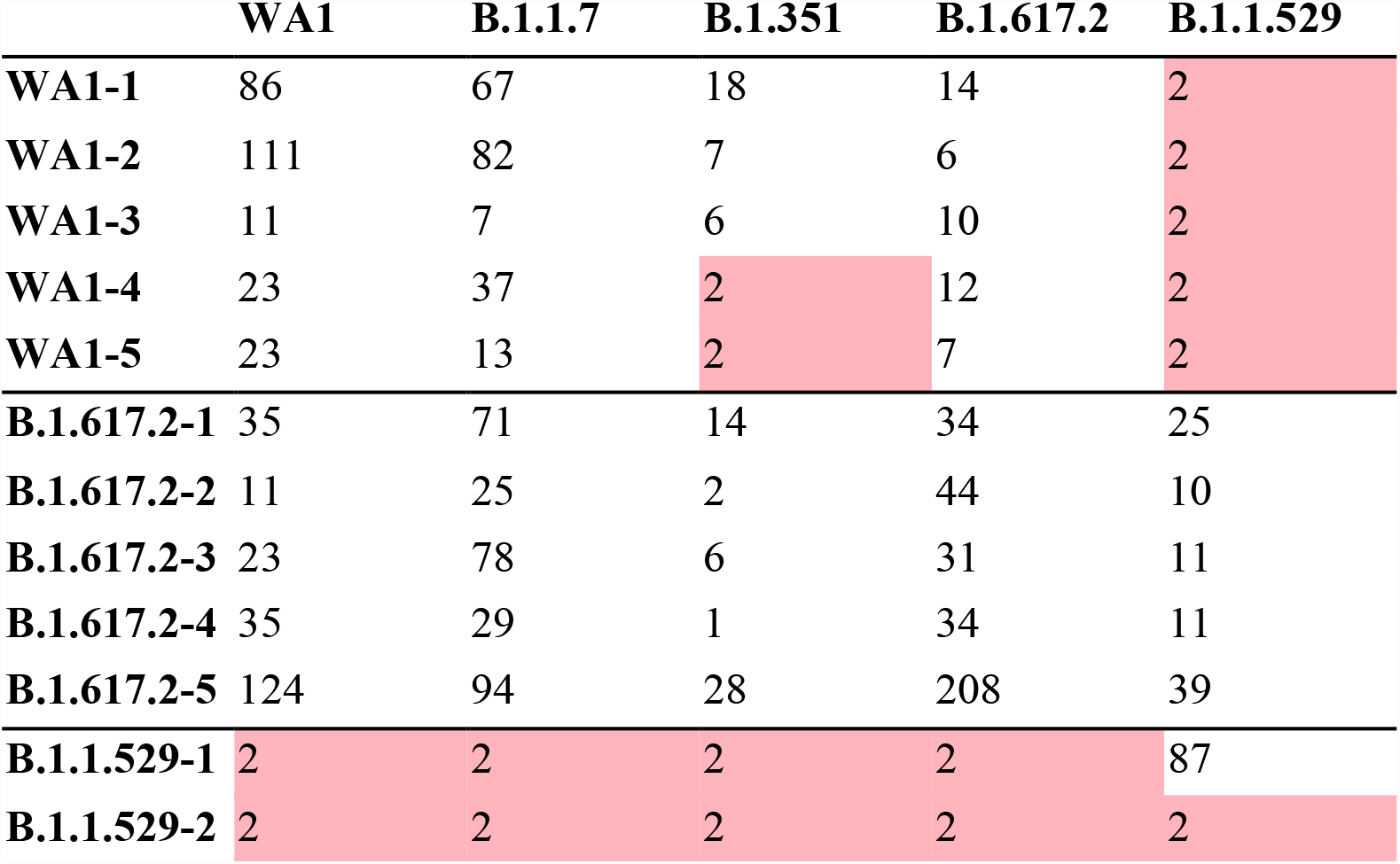
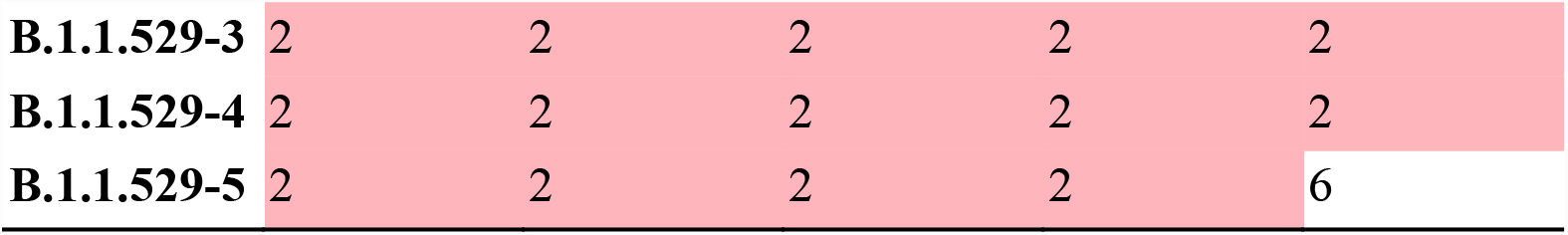
Neutralization titers for infected mice against VOCs.

**Fig. 4.**
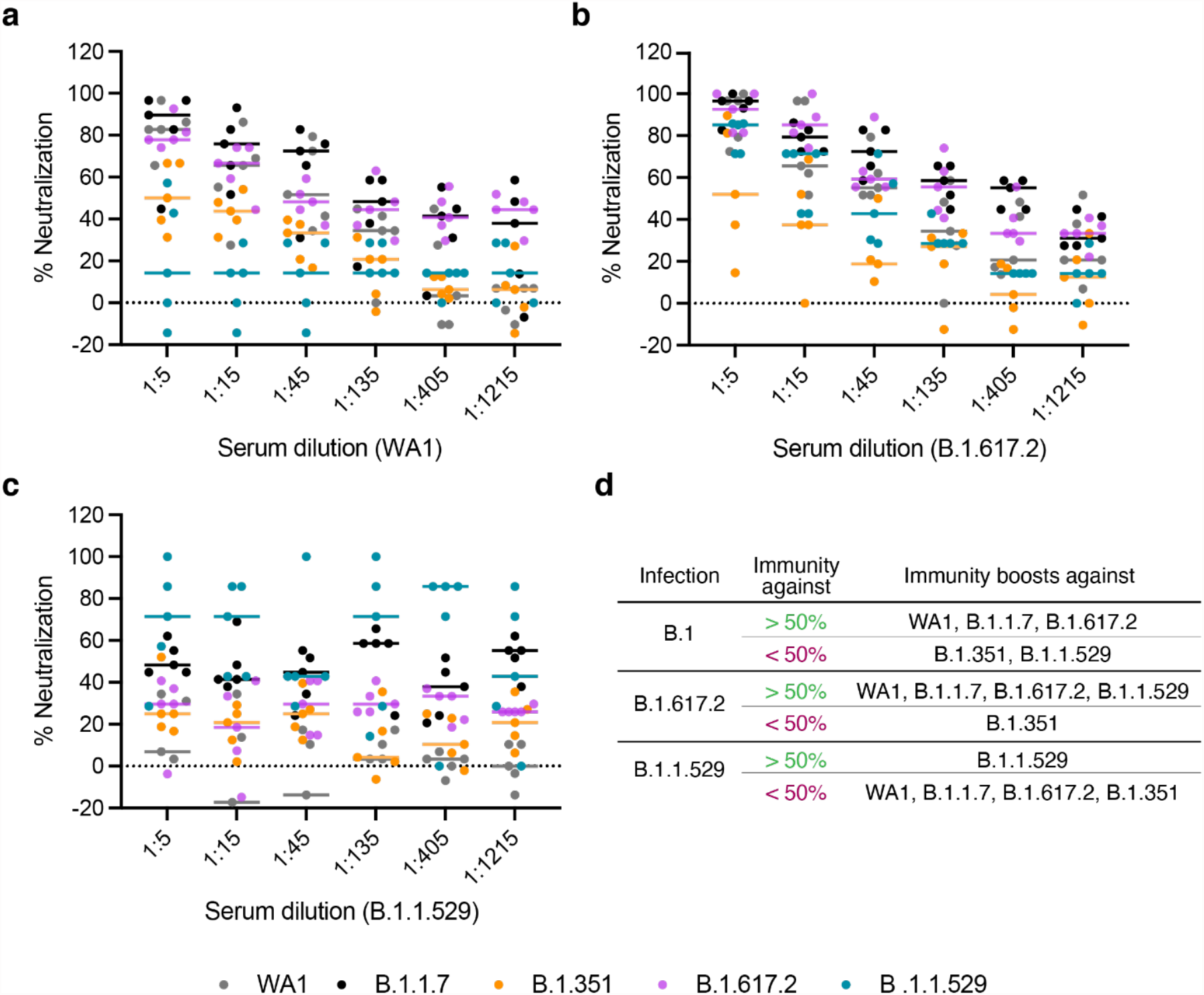
Cross-variant neutralization of SARS-CoV-2 isolates by sera from SARS-CoV-2 infected mice. **a**, Percent serum virus neutralization of SARS-CoV-2 isolates by sera from mice infected with WA1. Sera were taken at 7 dpi from mice (n=5) infected with respective virus isolates. The percent virus neutralization was expressed as PFU at indicated sera dilutions. **b**, Percent serum virus neutralization of SARS-CoV-2 isolates by sera from mice (n=5) infected with B.1.617.2. **c**, Percent serum virus neutralization of SARS-CoV-2 isolates by sera from mice (n=5) infected with B.1.1.529. **d**, Summary table representing immunity boost of mice sera evaluated in A-C, the median value at 1:5 and 1:15 serum dilutions indicating more than 50 % virus neutralization was considered as efficient whereas median less than 50% was considered as limited virus neutralization.

Next, we evaluated the degree of virus neutralizing immunity in convalescent sera (three to six weeks after infection) from individuals likely infected with Delta (collected during a local surge in July 2021). These sera effectively neutralized WA1 and Delta (>50%), but not Alpha, Beta, and Omicron strains (<50%). Naïve sera from uninfected and unvaccinated individuals did not show activity against any strain, as expected (**Fig. 5a,b,d, Table 2**). We then examined neutralizing activity elicited after breakthrough infections. Sera from likely Delta breakthrough cases (collected at two timepoints 10–17 days apart from July to November 2021) were tested in a virus-like particle (VLP) entry assay^23^. Neutralization was observed for all strains tested, albeit Omicron with a lower NT50, and increased significantly between the two time points for the ancestral and Delta strains (**Fig. 5e**). Notably, sera from vaccinated individuals with confirmed Omicron breakthrough infection showed the highest level of protection (>80%) against all strains, including Omicron (**Fig. 5c,d, Table 2**). These findings suggest that Omicron infection can effectively boost existing immunity conferred by the vaccination against other variants, eliciting “hybrid immunity” that is effective against not only itself but also other variants.

**Table 2.** Neutralization titers for naive, convalescent, vaccinated+B.1.1.529 individuals against VOCs.

|  | **WA1** | **B.1.1.7** | **B.1.351** | **B.1.617.2** | **B.1.1.529** |
| --- | --- | --- | --- | --- | --- |
| **CUR01** | 2 | 2 | 2 | 2 | 2 |
| **CUR02** | 2 | 2 | 2 | 2 | 2 |
| **CUR03** | 2 | 2 | 2 | 2 | 2 |
| **CUR04** | 2 | 2 | 2 | 2 | 2 |
| **CUR05** | 2 | 2 | 2 | 2 | 2 |
| **PC002** | 17 | 12 | 11 | 2 | 2 |
| **PC006** | 2 | 13 | 22 | 2 | 5 |
| **PC007** | 31 | 10 | 53 | 2 | 2 |
| **PC008** | 2 | 2 | 2 | 33 | 2 |
| **PC009** | 71 | 36 | 30 | 198 | 5 |
| **UMPIRE-1** | 3821 | 3241 | 626 | 723 | 407 |
| **UMPIRE-2** | 974 | 5942 | 618 | 437 | 40 |

**Fig. 5.**
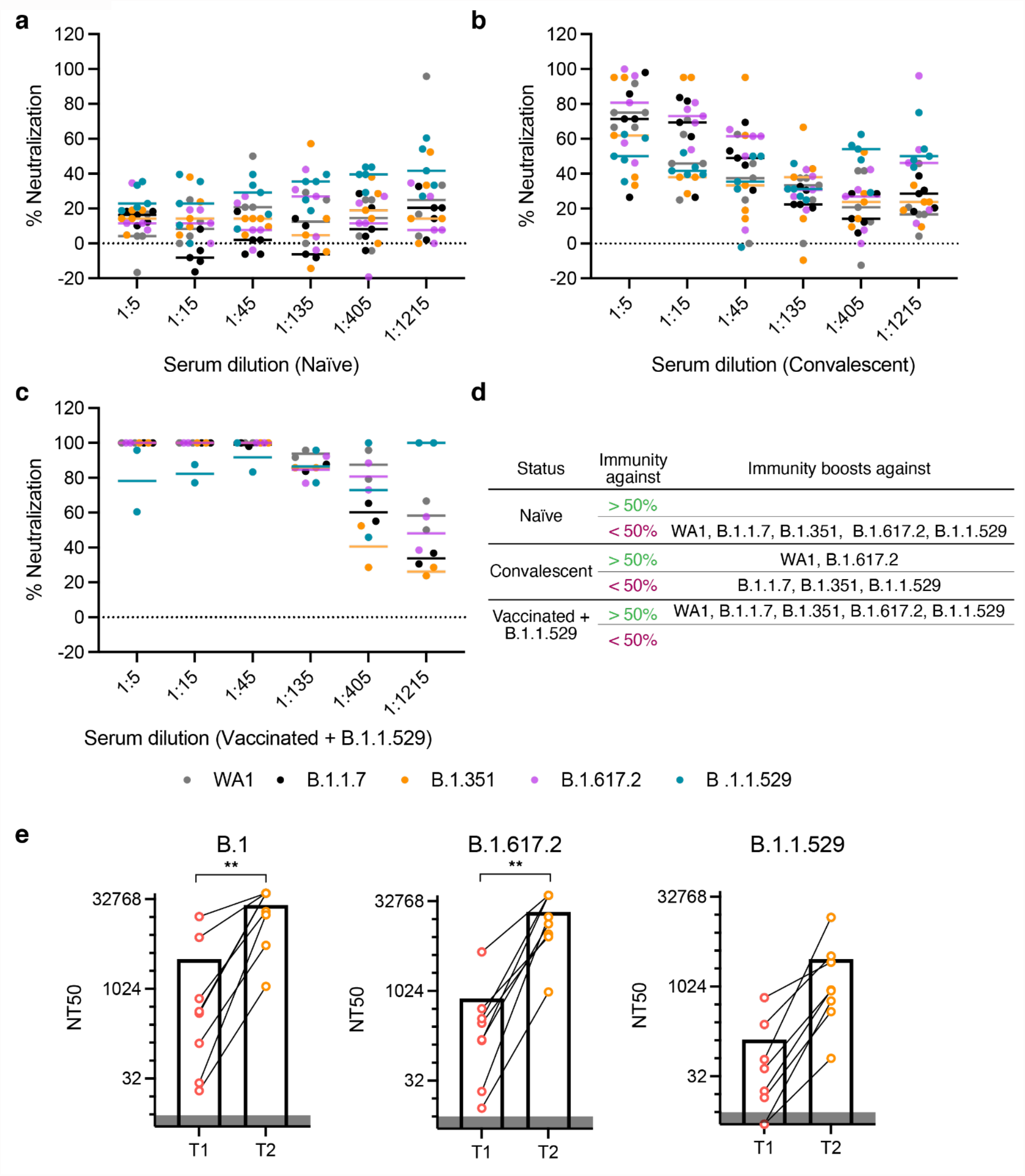
Cross-variant neutralization of SARS-CoV-2 isolates by human sera. **a**, Percent serum virus neutralization of SARS-CoV-2 isolates by sera from naïve (were never exposed to SARS-CoV-2 infection) individuals. **b**, Percent serum virus neutralization of SARS-CoV-2 isolates by sera from convalescent COVID-19 patients. **c**, Percent serum virus neutralization of SARS-CoV-2 isolates by sera from individuals vaccinated with breakthrough infection of B.1.1.529. **d**, Summary table representing immunity boost of sera evaluated in A-C. The median value at 1:5 and 1:15 serum dilutions indicating more than 50% virus neutralization was considered as efficient whereas median less than 50% was considered as limited virus neutralization. **e**, Serum neutralization of VLPs generated with different S genes from B.1, B.1.617.2, and B.1.1.529. 50% neutralization titers (NT50) of sera isolated from vaccinated individuals that had a breakthrough infection are shown and analyzed by student’s t test. **p<0.001.

Collectively, our study shows that while the Omicron virus is immunogenic, infection with this variant in unvaccinated individuals may not elicit effective cross-neutralizing antibodies against other variants. In vaccinated individuals, however, Omicron infection effectively induces immunity against itself and enhances protection against other variants. This, together with our finding that Delta infection is broadly immunogenic in mice, supports the inclusion of Omicron- and Delta-based immunogens in future multivalent vaccination strategies for broad protection against variants.

## Data Availability

All data produced in the present work are contained in the manuscript

## Methods

### Cell Culture

#### Human Lung Organoids

Whole human lung tissue was digested to a single cell suspension and plated in basement membrane extract as previously published^24^. Briefly, organoids were maintained in DMEM supplemented with supplemented with 10% (vol/vol) R-spondin1 conditioned media, 1% B27 (Gibco), 25 ng/mL Noggin (Peprotech), 1.25 mM N-Acetylcysteine (Sigma-Aldrich), 10 mM Nicotinamide (Sigma-Aldrich), 5 nM Herefulin Beta-1 (Peprotech), and 100 µg/mL Primocin (InvivoGen). HAO media is further supplemented with 5 µM Y-27632, 500 nM A83-01, 500 nM SB202190, 25 ng/mL FGF-7, 100 ng/mL FGF-10 (all from Stem Cell Technologies), HAO media was replaced every 3-4 days.

A549 cells expressing ACE2 (A549-ACE2) and Vero cells expressing TMPRSS (Vero-TMPRSS2) were a gift from O. Schwartz and S.P.J. Whelan, respectively. A549-ACE2 and Vero-TMPRSS2 cells were cultured in DMEM supplemented with 10% FBS and blasticidin (20ug/ml) (Sigma) at 37°C and 5% CO_2_. Short Terminal Repeat (STR) analysis by the Berkeley Cell Culture Facility on 17 July 2020 authenticates these as A549 cells with 100% probability.

Vero stably co-expressing human ACE2 and TMPRSS2 cells (gifted from A. Creanga and B. Graham at NIH) were maintained at 37°C and 5% CO2 in Dulbecco’s Modified Eagle medium (DMEM; Gibco) supplemented with 10% fetal calf serum, 100ug/mL penicillin and streptomycin (Gibco) and 10μg/mL of puromycin (Gibco).

293T cells stably co-expressing ACE2 and TMPRSS2 were generated through sequential transduction of 293T cells with TMPRSS2-encoding (generated using Addgene plasmid #170390, a gift from Nir Hacohen and ACE2-encoding (generated using Addgene plasmid #154981, a gift from Sonja Best) lentiviruses and selection with hygromycin (250 µg/mL) and blasticidin (10 µg/mL) for 10 days, respectively. ACE2 and TMPRSS2 expression was verified by western blot.

#### SARS-CoV-2 virus culture

SARS-CoV-2/human/USA/USA-WA1/2020 (WA1) (BEI NR-52281), B.1.1.7 (California Department of Health), B.1.351 (BEI NR-54008), B.1.617.2 (BEI NR-55611) and B.1.1.529 (California Department of Health) were used for either animal infection studies, or serum virus neutralization. The virus infection experiments were performed in a Biosafety Level 3 laboratory. Working stocks of SARS-CoV-2 were made in Vero-TMPRSS2 cells and were stored at -80°C until used.

Omicron variant was isolated from a nasopharyngeal swab sample from a patient hospitalized with COVID-19 at UCSF. A 200 uL aliquot of the sample was serially diluted 1:1 with media (DMEM supplemented with 1x penicillin/streptomycin) in a 96-well plate for 5 dilutions, in duplicate. A 100 uL of freshly trypsinized Vero-hACE2-TMPRSS2 cells, resuspended in infection media (made as above but with 2x penicillin/streptomycin, 5ug/mL amphotericin B [Bioworld] and no puromycin) were added to the nasal sample dilutions at 2.5×105 cells/mL concentration. Cells were cultured at 37°C and 5% CO_2_ and checked for cytopathic effect (CPE) from day 2-3. Vero-hACE2-TMPRSS2 cells form characteristic syncytia upon infection with SARS-CoV-2, enabling rapid and specific visual evaluation for CPE. Supernatants were harvested on day 3 after inoculation. A 200ul aliquot of P0 was used to infect a confluent T25 flask to generate a P1 culture, harvested after 3 days. Virus stocks were titered by plaque assay and sequence confirmed by nanopore sequencing.

#### K18-hACE2 mouse infection model

All protocols concerning animal use were approved (AN169239-01A) by the Institutional Animal Care and Use committees at the University of California, San Francisco and Gladstone Institutes and conducted in strict accordance with the National Institutes of Health Guide for the Care and Use of Laboratory Animal (Council, 2011). Mice were housed in a temperature- and humidity-controlled pathogen-free facility with 12-hour light/dark cycle and ad libitum access to water and standard laboratory rodent chow.

Briefly, the study involved intranasal infection of 6-8-week-old female K18-hACE2 mice with B.1.617.2 and B.1.1.529, while WA1 served as a control strain of SARS-CoV-2. A total of 15 animals were infected for each variant. Five mice from each group were euthanized at day two, four and seven post infection. The brain, upper respiratory tract including bronchus and nasal turbinates and lungs were processed for further analysis of virus replication.

#### Cellular infection studies

A549-ACE2 cells were seeded into 12-well plates. Cells were rested for at least 24 hours prior to infection. At the time of infection, media containing viral inoculum (MOI 0.01 and 0.1) was added on the cells. One hour after addition of inoculum, the media was replaced with fresh media without viral inoculum. The supernatant was harvested at 24, 48, and 72 hpi for further plaque assays.

#### Organoid infection studies

Organoids were plated on geltrex-coated plates (ThermoFisher, 12760013) with 100,000 cells per well, and infected at an MOI of 1. Two hours after addition of the inoculum, the supernatant was removed, cells were washed with PBS, and fresh HAO media was added. Supernatant was harvested for a plaque assay at 24 and 48 hours.

#### Virus neutralization assay

K18-hACE2 mice infected with WA1, B.1.617.2 and B.1.1.529 (n=5). Considering the early humane endpoints with WA1 and B.1.617.2, more animals (n=15) were infected for these groups, The serum samples from mice were collected at 7 dpi. Mock infected animals served as a control. The serum dilutions (50µL) were made as 1:5, 1:15, 1:45, 1:135, 1:405, 1:1215 in serum-free DMEM. The dilutions were separately added with 50 PFU (50µL) of SARS-CoV-2 WA1, B.1.1.7, B.1.351, B.1.617.2, B.1.1.529. The mixture was mixed gently, incubated at 37^°^C for 30 mins, followed by a plaque assay.

#### Plaque assays

Tissue homogenates and cell supernatants were analyzed for viral particle formation for *in vivo* and *in vitro* experiments, respectively. Briefly, Vero-TMPRSS2 were seeded and incubated overnight. The cells were inoculated with 10-1 to 10-6 dilutions of the respective homogenates or supernatant in serum-free DMEM. After the 1 hour absorption period, the media in the wells was overlaid with 2.5% Avicel (Dupont, RC-591). After 72 hours, the overlay was removed, the cells were fixed in 10% formalin for one hour, and stained with crystal violet for visualization of plaque forming units.

#### Quantitative polymerase chain reaction

RNA was extracted from cells, supernatants, or tissue homogenates using RNA-STAT-60 (AMSBIO, CS-110) and the Direct-Zol RNA Miniprep Kit (Zymo Research, R2052). RNA was then reverse-transcribed to cDNA with iScript cDNA Synthesis Kit (Bio-Rad, 1708890). qPCR reaction was performed with cDNA and SYBR Green Master Mix (Thermo Scientific) using the CFX384 Touch Real-Time PCR Detection System (Bio-Rad). See Table S1 for primers sequences. N gene standards were used to generate a standard curve for copy number quantification. N gene standard was generated by PCR using extracted genomic SARS-CoV-2 RNA as template. A single product was confirmed by gel electrophoresis and DNA was quantified by Nanodrop.

#### CyTOF analysis of mouse lung specimens

The mice used in the CyTOF study were infected with 5×102 PFU of WA1 and monitored for clinical signs of infection (e.g. body weight and body temperature) starting from day 1 to day 9 post infection. CyTOF was conducted similar to methods recently described25–28. Single-cell suspensions of lung tissue specimens processed using the GentleMACS system (Miltenyi) were treated with 25 μM cisplatin (Sigma) for 60 seconds as a viability dye. The cells were then quenched with CyFACS buffer (PBS supplemented with 0.1% BSA and 0.1% sodium azide) and fixed for 10 minutes with 2% paraformaldehyde (PFA; Electron Microscopy Sciences). Cells were then washed twice with CyFACs and frozen at −80°C until CyTOF antibody staining. Prior to antibody staining, specimens were barcoded using the Cell-ID TM 20-Plex PD Barcoding kit (Fluidigm, South San Francisco, CA, USA). Fc blocking was performed by treating the cells with 1.5% mouse and rat sera (both from Thermo Fisher) and 0.3% human AB sera (Sigma-Aldrich) for 15 minutes at 4°C. After washing with CyFACS, cells were stained for 45 minutes at 4°C with the cell surface antibodies listed in Extended Table 2. Antibodies were purchased pre-conjugated from Fluidigm, or conjugated using the MaxPAR conjugation kit (Fluidigm). After staining, cells were washed with CyFACS and fixed overnight at 4°C in 2% PFA, and then permeabilized for 30 minutes with Foxp3 Fix/Permeabilization Buffer (Fisher Scientific). After two washes with Permeabilization Buffer (Fisher Scientific), cells were Fc blocked again for 15 minutes at 4°C with mouse and rat sera diluted in Permeabilization Buffer. After washing with Permeabilization Buffer, cells were stained for 45 minutes at 4°C with the intracellular antibodies listed in Extended Table 2. Prior to CyTOF analysis, cells were incubated for 20 minutes with a 1:500 dilution DNA intercalator (Fluidigm), and then washed twice with CyFACS and once with Cell Acquisition Solution (CAS, Fluidigm). Acquisition was performed in the presence of EQ TM Four Element Calibration Beads (Fluidigm) diluted in CAS. Cells were analyzed on a CyTOF 2 instrument (Fluidigm) at the UCSF Parnassus Flow Core. For data analysis, CyTOF datasets were normalized to EQ calibration and manually gated using the FlowJO software (BD Biosciences). tSNE visualizations of the datasets were performed in Cytobank, with default settings.

#### VLP production

For a 6-well, plasmids CoV2-N (0.67), CoV2-M-IRES-E (0.33), CoV-2-Spike (0.0016) and Luc-T20 (1.0) at indicated mass ratios for a total of 4 µg of DNA were diluted in 200 µL optimem. 12 µg PEI was diluted in 200 µL Opti-MEM and added to plasmid dilution quickly to complex the DNA. Transfection mixture was incubated for 20 minutes at room temperature and then added dropwise to 293T cells in 2 mL of DMEM containing fetal bovine serum and penicillin/streptomycin. Media was changed after 24 hours of transfection and At 48 hours post-transfection, VLP containing supernatant was collected and filtered using a 0.45 µm syringe filter. For other culture sizes, the mass of DNA used was 1 µg for 24-well, 4 µg for 6-well, 20 µg for 10-cm plate and 60 µg for 15-cm plate. Optimem volumes were 100 µL, 400 µL, 1 mL and 3 mL respectively and PEI was always used at 3:1 mass ratio.

#### VLP luciferase assay

In each well of a clear 96-well plate 50 µL of SC2-VLP containing supernatant was added to 50 µL of cell suspension containing 50,000 receiver cells (293T ACE2/TMPRSS2). Cells were allowed to attach and take up VLPs overnight. Next day, supernatant was removed and cells were rinsed with 1X PBS and lysed in 20 µL passive lysis buffer (Promega) for 15 minutes at room temperature with gentle rocking. Lysates were transferred to an opaque white 96-well plate and 50 µL of reconstituted luciferase assay buffer was added and mixed with each lysate. Luminescence was measured immediately after mixing using a TECAN plate reader.

#### Human serum neutralization assay against VLPs

Human serum samples were acquired from two ongoing clinical trials led by Curative and UCSF or from hospitalized patients at UCSF. The Curative clinical trial protocol was approved by Advarra under Pro00054108 for a study designed to investigate immune escape by SARS-CoV-2 variant (University of California, Los Angeles Protocol Record PTL-2021-0007, ClinicalTrials.gov Identifier NCT05171803). Sample specimens were collected from adults (18-50 years) who either had been vaccinated for COVID-19 and/or had a history of COVID-19. Sample acquisition involved standard venipuncture procedure to collect a maximum of 15 ml whole blood, incubation at ambient temperature for 30–60 min to coagulate, centrifugation at 2200–2500 rpm for 15 min at room temperature, and storage on ice until delivered to the laboratory for serum aliquoting and storage at − 80 °C until use. A quantitative SARS-CoV-2 IgG ELISA was performed on serum specimens (EuroImmun, Anti-SARS-CoV-2 ELISA (IgG), 2606–9621G, New Jersey). Remnant plasma samples from patients hospitalized with COVID-19 at UCSF were obtained from UCSF Clinical Laboratories daily based on availability. Remnant samples were aliquoted and biobanked and retrospective medical chart review for relevant demographic and clinical metadata were performed under a waiver of consent and according to “no subject contact” protocols approved by the UCSF Institutional Review Board (protocol number 10-01116). Plasma samples were also collected through the UMPIRE (UCSF EMPloyee and community member Immune REsponse) study (protocol number 20-33083), a longitudinal COVID-19 research study focused on collection of prospective whole blood and plasma samples from enrolled subjects to evaluating the immune response to vaccination, with and without boosting, and/or vaccine breakthrough infection. The study cohorts included (1) fully vaccinated individuals with either 2 doses of Emergency Use Authorization (EUA) authorized mRNA vaccine (Pfizer or Moderna) or 1 dose of the EUA authorized Johnson and Johnson vaccine. Consented participants came to a UCSF CTSI Clinical Research Service (CRS) Laboratory where their blood was drawn by nurses and phlebotomists. At each visit, two to four 3mL EDTA tubes of whole blood were drawn, and one or two EDTA tubes were processed to plasma from each timepoint. Relevant demographic and clinical metadata from UMPIRE participants were obtained through participant Qualtric surveys performed at enrollment and at each blood draw. Serum samples were heat inactivated at 56°C for 30 mins prior to use in VLP or infection assays. Pre-COVID sera was pooled into one sample.

## Acknowledgements

We thank Stanley Tamaki and Claudia Bispo for CyTOF assistance at the Parnassus Flow Core, and the lab of Eliver Ghosn for guidance on lung cell processing. This research is funded by grants from the National Institutes of Health: NIAID R37AI083139 to M.O., NIH/NIAID (F31 AI164671-01) to I.P.C., NHLBI U54HL147127 to M.M. A.M.S is supported by Natural Sciences and Engineering Research Council of Canada (NSERC PDF-533021-2019). M.O. and W.C.G also received support from the Roddenberry Foundation and M.O. received a gift from Pam and Ed Taft. J.A.D. acknowledges support from the National Institutes of Health (R21AI59666) and support from the Howard Hughes Medical Institute and the Gladstone Institutes. N.R. acknowledges support from the Van Auken Private Foundation, David Henke, and Pamela and Edward Taft; and Awards #2164 and #2208 from Fast Grants, a part of Emergent Ventures at the Mercatus Center, George Mason University. C.Y.C thanks the staff at UCSF Clinical Laboratories and the UCSF Clinical Microbiology Laboratories for help in identifying and aliquoting nasal swab and plasma samples. CYC acknowledges support by the Innovative Genomics Institute (IGI) at UC Berkeley and UC San Francisco, US Centers for Disease Control and Prevention contract 75D30121C10991, Abbott Laboratories, and the Sandler Program for Breakthrough Biomedical Research at UCSF. The group also acknowledges support from the James B. Pendleton Charitable Trust. The funders had no role in study design, data collection and analysis, decision to publish, or preparation of the manuscript.

## Author Contributions

Conceptualization: RKS, IPC, MO

Investigation: RKS, IPC, TM, AMS, CRS, AC, MMK, BS, PC, AG

Anti sera: NB, PS, AS, VS, AG, JN, IS, BM, NK, VH, MS, LL, MB, FT, FWS, CYC, LS

Omicron virus culture: MAG, MKM, DW, CH, RA

Lung tissue for organoids: XF, MM, MM

BSL3 facility maintenance: MM Supervision: LS, JAD, NR, CYC, MO

Writing - original draft: RKS, IPC, MO

Writing - reviewing, & editing: RKS, IPC, GRK, WCG, TM, NR, MO

## Competing Interests

A.M.S. and J.A.D. are inventors on a patent application filed by the Gladstone Institutes and the University of California that covers the method and composition of SARS-CoV-2 VLP preparations for RNA transduction and expression in cells. J.A.D. is a cofounder of Caribou Biosciences, Editas Medicine, Scribe Therapeutics, Intellia Therapeutics, and Mammoth Biosciences. J.A.D. is a scientific advisory board member of Vertex, Caribou Biosciences, Intellia Therapeutics, eFFECTOR Therapeutics, Scribe Therapeutics, Mammoth Biosciences, Synthego, Algen Biotechnologies, Felix Biosciences, The Column Group, and Inari. J.A.D. is a director at Johnson & Johnson and Tempus and has research projects sponsored by Biogen, Pfizer, AppleTree Partners, and Roche. C.Y.C. is the director of the UCSF-Abbott Viral Diagnostics and Discovery Study and receives research support from Abbott Laboratories. C.Y.C. also receives support for SARS-CoV-2 research unrelated to this study from Mammoth Biosciences.

**Extended Table 1.**
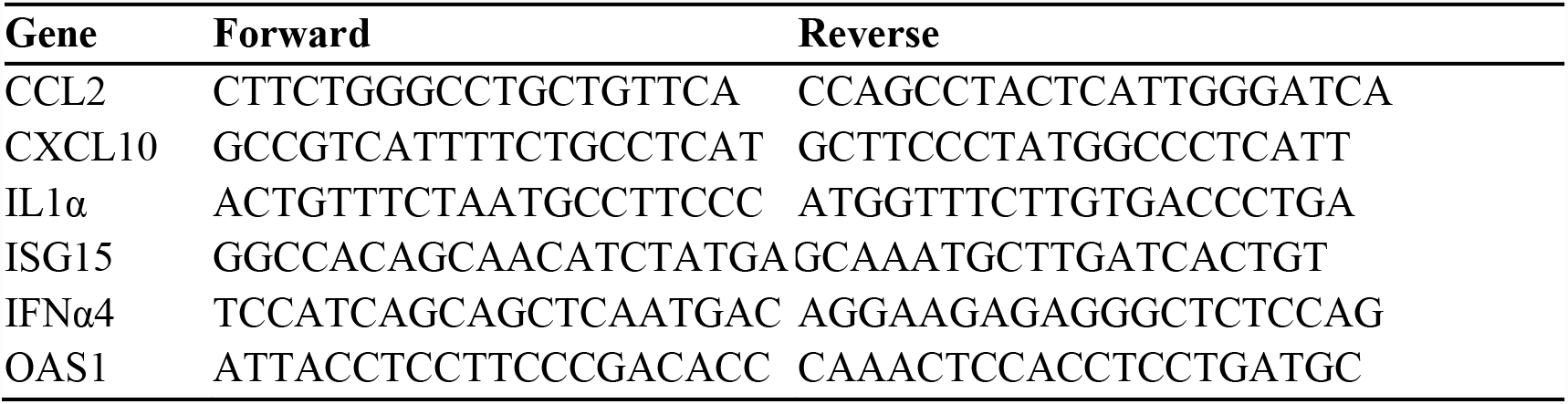
List of qPCR primers for mouse.

**Extended Table 2.** List of CyTOF staining antibodies for mouse.

| Antibody | Clone | Metal label | Vendor |
| --- | --- | --- | --- |
| Ly6G/C [Gr1] | RB6-8C5 | 141Pr | Fluidigm |
| CD11c | N418 | 142Nd | Fluidigm |
| CD69 | H1.2F3 | 145Nd | In-house |
| CD103 | 2E7 | 146Nd | In-house |
| CD45 | 30-F11 | 147Sm | Fluidigm |
| CD11b [MAC1] | M1/70 | 148Nd | Fluidigm |
| CD19 | 6D5 | 149Sm | Fluidigm |
| CD123 | 5B11 | 150Nd | In-house |
| CD25 | 3C7 | 151Eu | In-house |
| CD3e | 145-2C11 | 152Sm | Fluidigm |
| TER-119 | TER119 | 154Sm | Fluidigm |
| CXCR4 | L276F12 | 159Tb | In-house |
| CD62L | MEL-14 | 160Gd | In-house |
| CD127 | A7R34 | 161Dy | In-house |
| CXCR5 | L138D7 | 163Dy | In-house |
| PD1 | RMP1-30 | 164Dy | In-house |
| CD8α | 53-6.7 | 168Er | Fluidigm |
| TCRβ | H57-597 | 169Tm | Fluidigm |
| NK1.1 | PK136 | 170Er | Fluidigm |
| CD44 | IM7 | 171Yb | Fluidigm |
| CD4 | RM4-5 | 172Yb | Fluidigm |
| CD304 | 3E12 | 173Yb | In-house |
| B220 | RA3-6B2 | 176Yb | Fluidigm |
| IL-5<sup>#</sup> | TRFK5 | 143Nd | Fluidigm |
| IL-2<sup>#</sup> | JES6-5H4 | 144Nd | Fluidigm |
| IL21<sup>#</sup> | 149204 | 155Gd | In-house |
| Foxp3<sup>#</sup> | 3G3 | 156Gd | In-house |
| CTLA4<sup>#</sup> | UC10-4B9 | 157Gd | In-house |
| IL-10<sup>#</sup> | JES5-16E3 | 158Gd | Fluidigm |
| TNFα<sup>#</sup> | MP6-XT22 | 162Dy | Fluidigm |
| IFNγ<sup>#</sup> | XMG1.2 | 165Ho | Fluidigm |
| IL-4<sup>#</sup> | 11B11 | 166Er | Fluidigm |
| IL-6<sup>#</sup> | MP5-20F3 | 167Er | Fluidigm |
| IL-17A<sup>#</sup> | TC11-18H10.1 | 174Yb | Fluidigm |
| IFNα<sup>#</sup> | F18 | 175Lu | In-house |

<sup>#</sup>*Intracellular antibodies*

## Figure Legends

**Extended Data 1.**
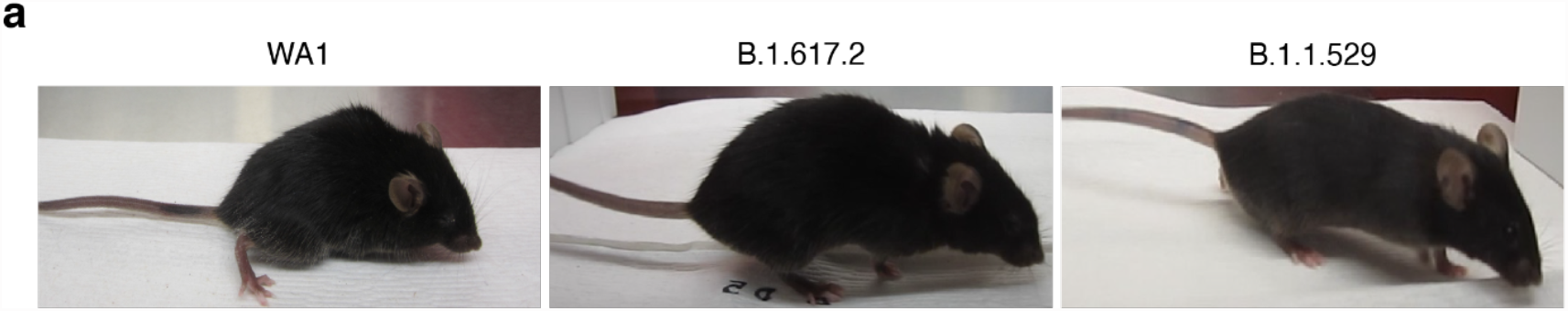
Physical conditions of the infection mice at 5 dpi. **a**, Representative images of WA1-, B.1.617.2-, and B.1.1.529-infected mice 5 dpi. WA1-infected mice were lethargic and had a hunched posture, ungroomed coat, and squinted eyes. B.1.617.2-infected mice are mildly lethargic. B.1.1.529-infected mice appeared normal.

**Extended Data 2.**
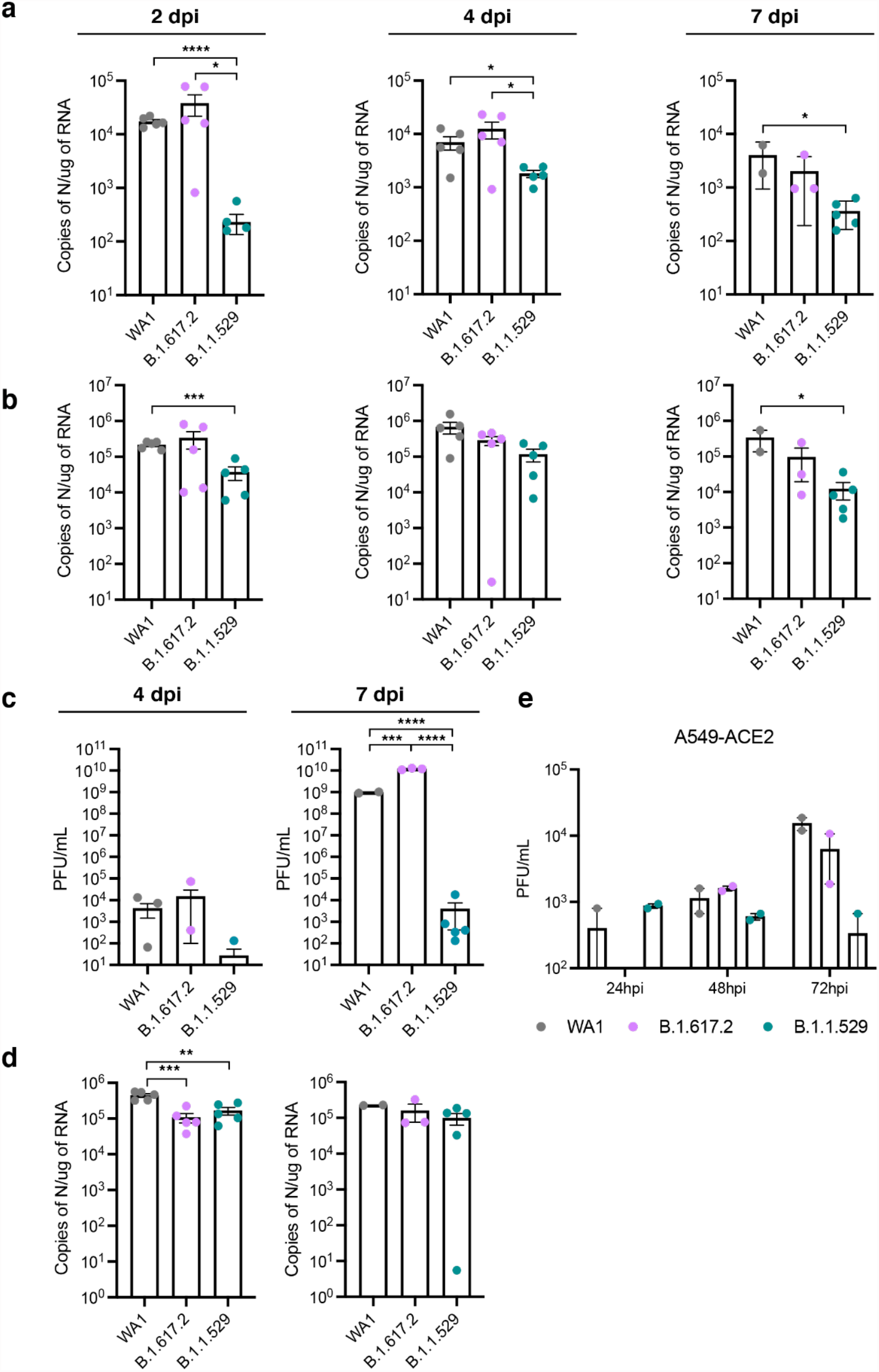
Lower viral replication of Omicron in mice and human cells. **a**, RT-qPCR of SARS-CoV-2 N RNA isolated from upper respiratory tract (nasal turbinates and bronchus) of WA1 (grey), B.1.617.2 (purple), and B.1.1.529 (teal) infected mice at indicated time points. Data are expressed in absolute copies/ug based on a standard curve of N gene with known copy number. Data is shown as an average ± SEM at each time point and analyzed by student’s t-test. *p<0.05, ****p<0.0001. **b**, RT-qPCR of SARS-CoV-2 N RNA isolated from lungs of infected mice at indicated time points. Data are expressed in absolute copies/ug based on a standard curve of N gene with known copy number. Data is shown as an average ± SEM at each time point and analyzed by the student’s t-test. *p<0.05, ***p<0.0005. **c**, Plaque assay titers from the brains of infected mice at indicated time point. Data is shown as an average ± SEM at each time point and analyzed by the student’s t-test. ***p<0.001, ****p< 0.0001. **d**, RT-qPCR of SARS-CoV-2 N RNA isolated from brains of infected mice at indicated time points. Data are expressed in absolute copies/ug based on a standard curve of N gene with known copy number. Data is shown as an average ± SEM at each time point and analyzed by the student’s t-test. **p<0.01, ***p<0.001. **d**, Plaque assay titers from supernatants of infected A549-ACE2 (MOI of 0.01). Data is shown as average ± SEM.

**Extended Data 3.**
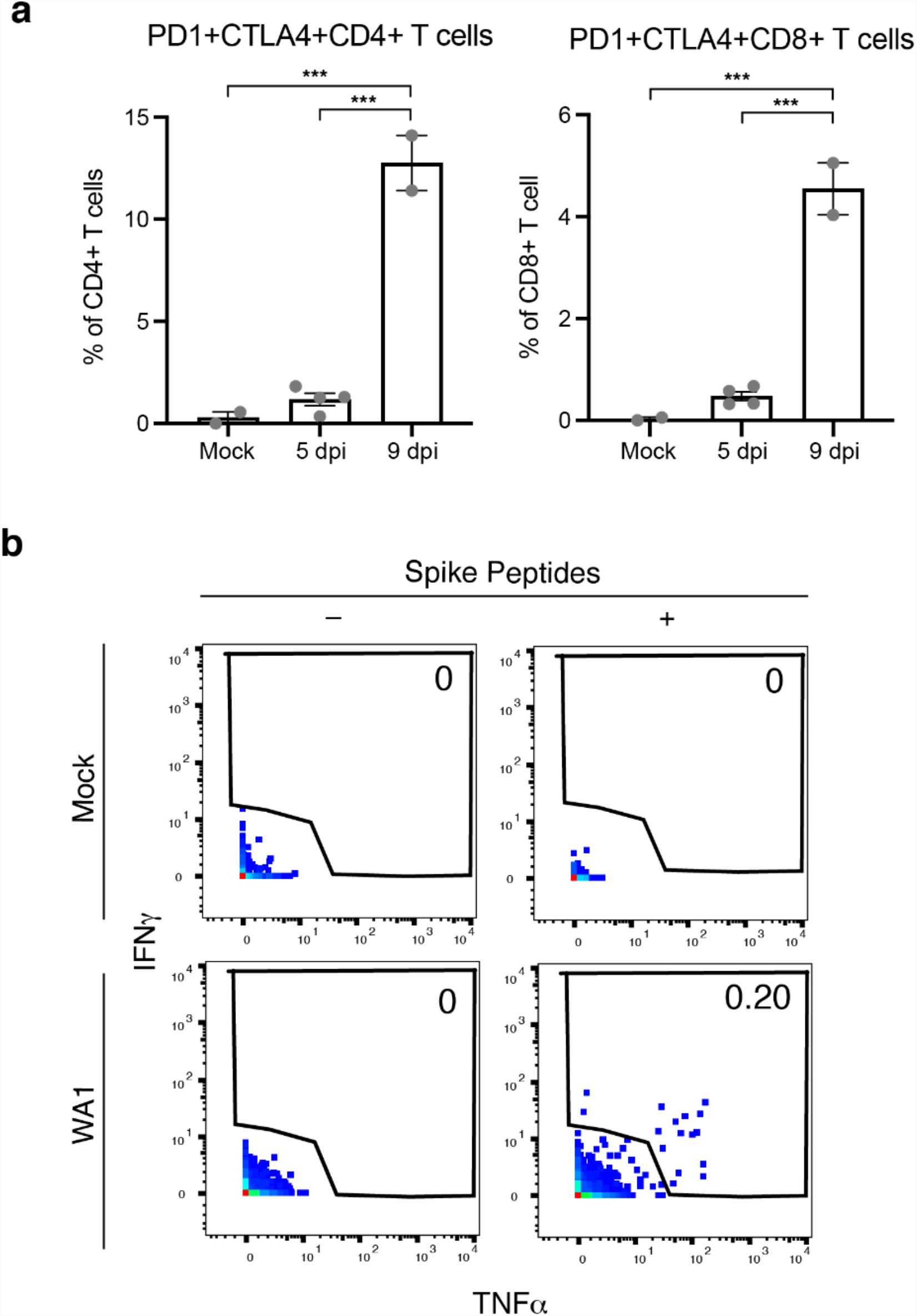
SARS-CoV-2 specific T cells. **a**, SARS-CoV-2-specific T cells are elicited in lungs of WA1-infected mice. Representative plots corresponding to pulmonary T cells from mock infected (left) and WA1-infected (right) K18-hACE2 mice, stimulated for 6 hours with or without overlapping SARS-CoV-2 spike peptides. Note-SARS-CoV-2-specific T cells (those producing IFNγ and/or TNFα) were only detected in infected mice after peptide stimulation. **b**, T cells from lungs of infected mice harbor significantly higher numbers of PD1+CTLA4+ T cells. ***p<0.001 as assessed using a one-way ANOVA and adjusted for multiple testing using the Bonferroni.

